# Use of estimands in cluster randomised trials: a review

**DOI:** 10.1101/2025.06.10.25329251

**Authors:** Dongquan Bi, Andrew Copas, Brennan C Kahan

## Abstract

**Background:** An estimand is a clear description of the treatment effect a study aims to quantify. The ICH E9(R1) addendum lists five attributes that should be described as part of the estimand definition. However, the addendum was primarily developed for individually randomised trials. Cluster randomised trials (CRTs), in which groups of individuals are randomised, have additional considerations for defining estimands (e.g., how individuals and clusters are weighted, how cluster-level intercurrent events are handled, etc). However, it is currently unknown if estimands are being used in CRTs, or whether the considerations specific to CRTs are being described.

**Methods:** We reviewed 73 CRTs published between Oct 2023 and Jan 2024 that were indexed in MEDLINE. For each trial, we assessed whether the estimand for the primary outcome was described, or if not, whether it could be inferred from the statistical methods. We also assessed whether considerations specific to CRTs were described or inferable, how trials were analysed, and whether key assumptions being made in the analysis (e.g. “no informative cluster size”) could be identified.

**Results:** No trials attempted to describe the estimand for their primary outcome. We were able to infer the full estimand based on the five attributes outlined in ICH E9(R1) in only 49% of trials, and when including additional considerations specific to CRTs, this figure dropped to 21%. Key drivers of this ambiguity were lack of clarity around whether individual- or cluster-average effects were of interest (unclear in 63% of trials), and how cluster-level intercurrent events were handled (unclear in 21% of trials for which this was applicable). Over half of trials used mixed-effects models or GEEs with an exchangeable correlation structure, which make the assumption that there is no informative cluster size; however, only one of these trials performed sensitivity analyses to evaluate robustness of results to deviations from this assumption. There were 14% of trials that used independence estimating equations or the analysis of cluster level summaries; however, because no trials stated whether they were targeting the individual- or cluster-average effect, it was impossible to determine whether these methods implemented the appropriate weighting scheme and were thus unbiased.

**Conclusion:** The uptake of estimands in published cluster randomised trial articles is low, making it difficult to ascertain which questions were being investigated or whether statistical estimators were appropriate for those questions. This highlights an urgent need to develop guidelines on defining estimands that cover unique aspects of CRTs to ensure clarity of research questions in these trials.

## Introduction

An estimand is a clear description of the treatment effect a study aims to quantify ^1, 2^. Estimands can help to both clarify the question a study sets out to investigate and ensure that appropriate statistical methods (estimators) are used to answer that question. In 2019 the ICH E9(R1) addendum introduced a structured approach to defining estimands, comprising of the specification of 5 attributes: (i) population of patients, (ii) treatment conditions being compared, (iii) the endpoint, (iv) population-level summary measure, and (v) how intercurrent events (post randomisation events that affect interpretation or existence of outcomes, such as treatment non-adherence) are handled.^1^

However, the ICH E9(R1) addendum was developed by medicines regulators in conjunction with the pharmaceutical industry, and as such was primarily developed for individually randomised trials. Cluster randomised trials (CRTs), where groups of individuals are randomised, may require specification of additional attributes. For example, investigators also need to consider the population of clusters for which they wish to estimate the treatment effect ^3–7^; how individuals and clusters are weighted (e.g. individual-vs. cluster-average effect) ^5, 7–12^; whether marginal or cluster-specific effects are of interest ^5, 11, 13, 14^; and how intercurrent events that occur at the cluster-level (such as if a cluster decided not to implement the intervention) are handled ^5^. Table 1 provides a summary of some additional considerations.

**Table 1.**
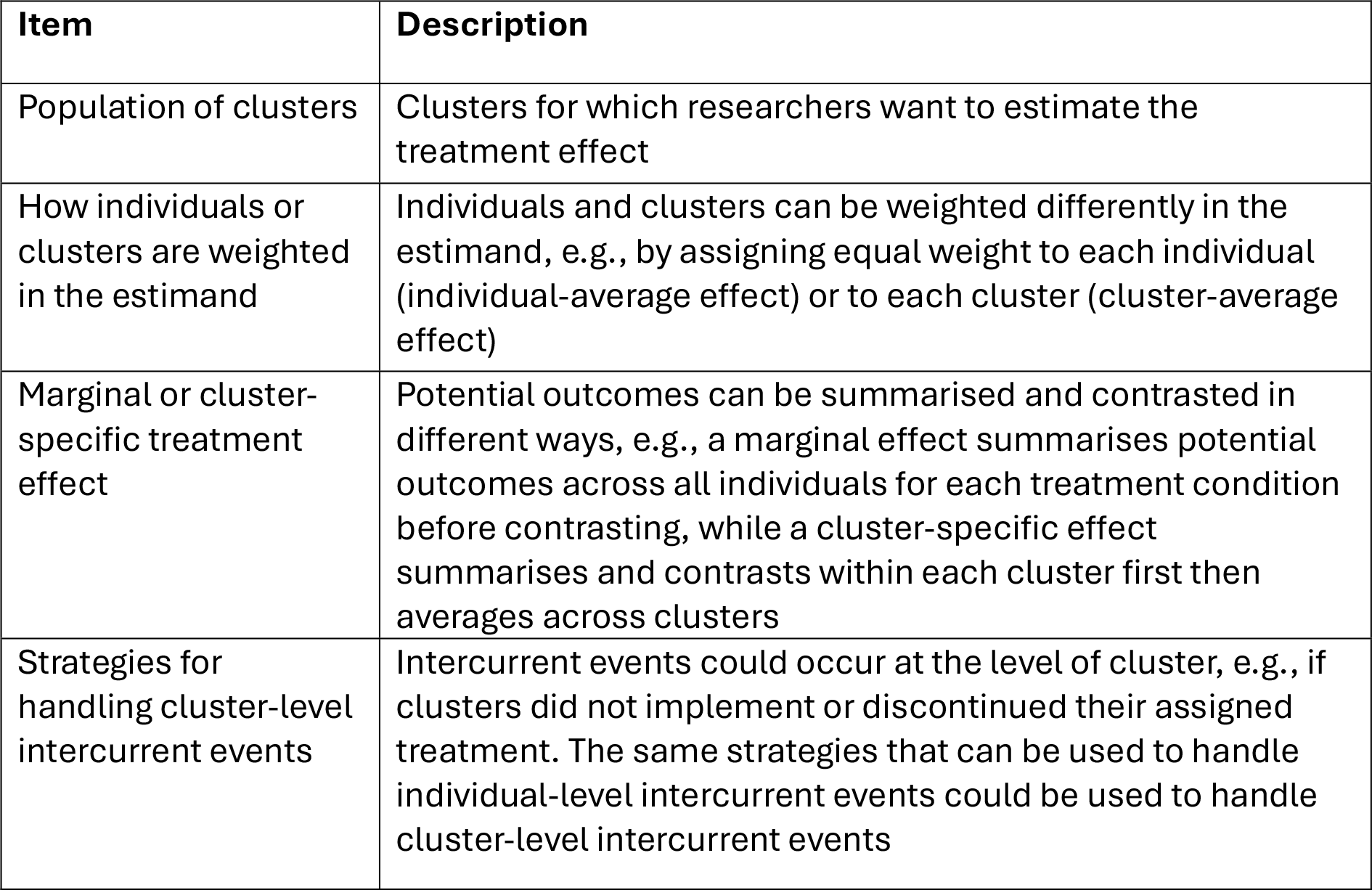
Summary of some additional considerations for defining estimands in cluster randomised trials.

| Item | Description |
| --- | --- |
| Population of clusters | Clusters for which researchers want to estimate the treatment effect |
| How individuals or clusters are weighted in the estimand | Individuals and clusters can be weighted differently in the estimand, e.g., by assigning equal weight to each individual (individual-average effect) or to each cluster (cluster-average effect) |
| Marginal or cluster-specific treatment effect | Potential outcomes can be summarised and contrasted in different ways, e.g., a marginal effect summarises potential outcomes across all individuals for each treatment condition before contrasting, while a cluster-specific effect summarises and contrasts within each cluster first then averages across clusters |
| Strategies for handling cluster-level intercurrent events | Intercurrent events could occur at the level of cluster, e.g., if clusters did not implement or discontinued their assigned treatment. The same strategies that can be used to handle individual-level intercurrent events could be used to handle cluster-level intercurrent events |

Failure to take these additional considerations into account when defining estimands for CRTs can both create ambiguity around trial objectives, as well as hamper appraisal around the choice of estimator. For instance, common estimators for CRTs include generalising estimating equations (GEEs), mixed-effects models, independence estimating equations (IEEs), and the analysis of cluster-level summaries. Each estimator requires certain assumptions in order to be unbiased for a specific estimand. For instance, when there is informative cluster size (i.e. when outcomes or treatment effects vary according to cluster size), IEEs and the analysis of cluster-level summaries will only be unbiased if an appropriate weighting scheme is used which is aligned to the target estimand (e.g. individual- or cluster-average).^9^ Furthermore, under informative cluster size, both GEEs with an exchangeable correlation structure (termed “GEEs(exch)” hereafter) and mixed-effect models may be biased for both the individual- and cluster-average effects.^8, 15–17^ Without precise definition of the estimand including aspects unique to CRTs, it is impossible to know whether a trial’s estimator is aligned to its overall objective, or what assumptions are being made.

Despite growing recognition around the importance of estimands, current reporting guidelines for CRTs were established before the introduction of the ICH E9(R1) addendum ^7^, and it is currently unclear if estimands are being used in CRTs. Furthermore, due to lack of specific guidance on defining estimands in CRTs, even if estimands are being used, it is unclear whether studies are incorporating the considerations specific to CRTs. We therefore undertook a review of published CRTs to determine how often estimands are used, whether considerations specific to CRTs are being described, and whether the key assumptions of the chosen estimators could be identified and evaluated.

## Method

### Search strategy

We searched on 23^rd^ Jan 2024 for articles reporting results from a CRT published between 1^st^ Oct 2023 and 15^th^ Jan 2024 on MEDLINE. The full search strategy is available in Appendix 1 in the Supplemental Material. Briefly, the search strategy contained terms to (i) select a randomised controlled design, such as with publication type of “randomised controlled trial” or with keyword “random” in the abstract; (ii) select a cluster design, such as with the MeSH term “cluster random”; and (iii) exclude ineligible articles (described below), such as with publication type of “review” or with keyword “protocol” in the title or abstract.

### Eligibility

Parallel-group CRTs were eligible with no restrictions on medical conditions or type of intervention. Crossover, stepped wedge, and factorial designs were excluded owing to potential differences in estimands and statistical estimation considerations for these trials. Other exclusion criteria were pilot or feasibility studies, non-randomised studies, secondary analyses or a follow-up of a previously published trial, CRTs with cost effectiveness as the primary outcome, articles with more than one trial reported, meta-analyses, systematic reviews, interim analyses, and letters to the editor or commentaries.

Title and abstract screening for eligibility was performed by a single reviewer (DB). Queries regarding eligibility were discussed with at least one other author (BCK and/or AC).

### Data extraction

Data were extracted using a piloted standardised extraction form. The extraction form was built on the Qualtrics Platform. The full extraction form is available in Appendix 2 in the Supplemental Material. One author (DB) extracted data for all eligible articles. Another author (BCK) independently checked all extractions against the source article; discrepancies were resolved by discussion.

Extracted data included: trial characteristics, whether the estimand was described, and if not, whether it could be inferred from the statistical methods, the types of intercurrent events that occurred, the type of estimand that was used, and the statistical methods. Data on the estimand and statistical methods were extracted for the primary estimand for the trial’s primary outcome. Rules for determining the trial’s primary outcome and primary estimand are given in Appendix 3 in the Supplemental Material.

We evaluated whether certain types of intercurrent events were applicable. An intercurrent event was deemed applicable if the manuscript indicated it had occurred during the trial, or was considered during the trial’s planning stages, for instance if it was (i) reported as having occurred during the trial; (ii) reported as part of the estimand definition; or (iii) mentioned as part of the analysis strategy.

We evaluated whether each of the five standard estimand attributes outlined in the ICH E9(R1) addendum was “stated”, “inferable”, or “not inferable” using methods similar to those used in other recent reviews of estimands in individually randomised trials.^18, 19^ We also evaluated whether the additional considerations specific to CRTs listed in Table 1 were “stated”, “inferable”, or “not inferable”. Rules for determining whether attributes were stated, inferable, or not inferable are given in Appendix 4 in the Supplemental Material.

### Statistical methods

Data was summarised descriptively using frequencies and percentages. All analyses were performed using STATA version 18.

## Results

### Search results and trial characteristics

The search identified 192 articles, of which 73 were eligible (Figure 1). The 73 eligible articles were published between 01 Oct 2023 and 15 Jan 2024. Most trials had two treatment arms (86%) and used a psychological, behavioural, or education intervention (89%). The median sample size was 754 participants (IQR: 101, 3414) and 25 clusters (IQR: 8, 203). Further trial characteristics are summarised in Appendix 5 in the Supplemental Material.

**Figure 1.**
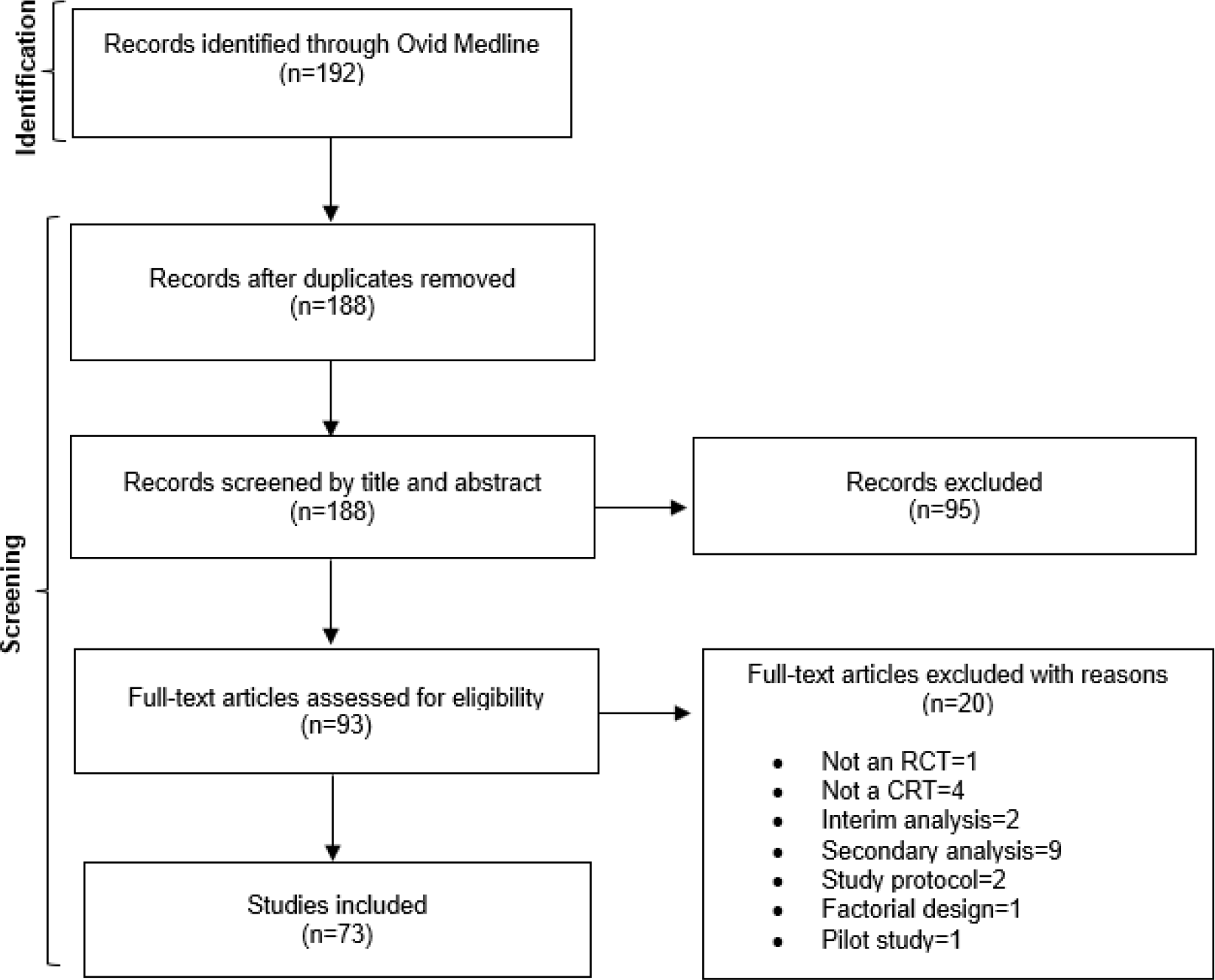
Flow diagram of the search process.

### Primary estimands

No trials described the estimand for their primary outcome (Table 2). Nevertheless, we were able to infer the full estimand based on the five standard attributes from the ICH E9(R1) addendum in 36 trials (49%). However, when including the additional considerations specific to CRTs, we were only able infer the full estimand in 15 trials (21%).

**Table 2.** How well estimands are described in cluster randomised trials.

| **Characteristic** | **Trials (N=73)** |
| --- | --- |
| Formally tried to describe estimand | 0 (0%) |
| **Standard estimand attributes<sup>a</sup>** |  |
| Population of patients |  |
| Inferable | 46 (63%) |
| Not inferable | 27 (37%) |
| Treatment conditions |  |
| Inferable | 51 (70%) |
| Not inferable | 22 (30%) |
| Endpoint |  |
| Inferable | 73 (100%) |
| Not inferable | 0 (0%) |
| Summary measure |  |
| Inferable | 60 (82%) |
| Not inferable | 13 (18%) |
| Strategies for handling of individual-level intercurrent events<sup>b</sup> |  |
| Inferable | 22 (43%) |
| Not inferable | 29 (57%) |
| **Additional considerations specific to CRTs<sup>a</sup>** |  |
| Population of clusters |  |
| Inferable | 61 (84%) |
| Not inferable | 12 (16%) |
| Strategies for handling of cluster-level intercurrent events<sup>c</sup> |  |
| Inferable | 26 (79%) |
| Not inferable | 7 (21%) |
| How participants are weighted (individual- vs. cluster-average effect) |  |
| Inferable | 27 (37%) |
| Not inferable | 46 (63%) |
| Marginal vs. cluster-specific effect |  |
| Inferable | 70 (96%) |
| Not inferable | 3 (4%) |
| Overall estimand with standard 5 attributes<sup>d</sup> |  |
| Inferable | 36 (49%) |
| Not inferable | 37 (51%) |
| Overall estimand including additional considerations<sup>e</sup> |  |
| Inferable | 15 (21%) |
| Not inferable | 58 (79%) |

- a. No studies stated any attribute of their estimand. - b. Denominator = 51; for 22 trials this was not applicable. - c. Denominator = 33; for 40 trials this was not applicable. - d. Population of patients, treatment conditions, endpoint, population-level summary, and strategies for handling individual-level intercurrent events. - e. Five attributes from ICH E9 (R1) addendum and additional considerations including population of clusters, strategies for handling cluster-level intercurrent events, individual-vs. cluster-average effect, marginal vs. cluster-specific effect.

A key driver of the ambiguity in the estimand when including the CRT-specific considerations was lack of clarity on whether the effect of interest was the individual-average or cluster-average treatment effect, as we were unable to infer this for 46 trials (63%). We were also unable to infer the population of clusters for 12 trials (16%), and how cluster-level intercurrent events were handled for 7 of the trials (21%) for which this was applicable (Table 2).

### Intercurrent events

Sixty-four trials (88%) reported at least one intercurrent event. Fifty-three trials (73%) reported at least one individual-level intercurrent event, and 32 trials (44%) reported at least one cluster-level intercurrent event.

None of the trials in which intercurrent events were applicable reported clearly stated the strategies they used to handle intercurrent events in their estimand (Table 2). Nevertheless, we were able to infer the strategies for handling individual-level intercurrent events in 22 trials (43%) and cluster-level intercurrent events in 26 trials (79%) (Table 2).

Treatment non-adherence/discontinuation was the most common intercurrent event; it was applicable at the individual-level in 38 trials (52%) and at cluster-level in 30 trials (41%) (Table 3).

**Table 3.** Whether handling strategies for intercurrent events were inferable or not.

| Intercurrent event | Trials n/N (%) |
| --- | --- |
| **Individual-level** |  |
| **Treatment non-adherence/discontinuation** |  |
| Applicable | 38/73 (52%) |
| Inferable | 24/38 (63%) |
| Not inferable | 14/38 (24%) |
| **Treatment switching** |  |
| Applicable | 4/73 (5%) |
| Inferable | 4/4 (100%) |
| **Mortality** |  |
| Applicable | 19/73 (26%) |
| Inferable | 5/19 (26%) |
| Not inferable | 14/19 (74%) |
| **Not starting treatment** |  |
| Applicable | 19/73 (26%) |
| Inferable | 10/19 (53%) |
| Not inferable | 9/19 (47%) |
| **Other intercurrent events<sup>a</sup>** |  |
| Applicable | 6/73 (8%) |
| Inferable | 3/6 (50%) |
| Not inferable | 3/6 (50%) |
| **Cluster-level** |  |
| **Treatment non-adherence/discontinuation** |  |
| Applicable | 30/73 (41%) |
| Inferable | 29/30 (97%) |
| Not inferable | 1/30 (3%) |
| **Not implementing treatment** |  |
| Applicable | 15/73 (21%) |
| Inferable | 9/15 (60%) |
| Not inferable | 6/15 (40%) |
| **Treatment switching** |  |
| Applicable | 5/73 (7%) |
| Inferable | 5/5 (100%) |
| **Other intercurrent events<sup>a</sup>** |  |
| Applicable | 7/73 (10%) |
| Inferable | 7/7 (100%) |

<sup>a</sup>. Full list of intercurrent events and the handling strategies can be found in Appendix 5.

In 31 trials (42%), we were able to infer how at least one type of intercurrent event was handled (Table 4). Treatment policy was the most common strategy and was used to handle all individual-level intercurrent events in 26 trials (84%) and all cluster-level intercurrent events in 30 trials (97%). Five trials (16%) used a hypothetical strategy to handle at least one individual-level intercurrent event, and 1 trial (3%) used a composite strategy to handle at least one cluster-level intercurrent event.

**Table 4.** Strategies used to handle intercurrent events.

| Item |  |
| --- | --- |
| **By type of intercurrent event** | **n/N<sup>a</sup> (%)** |
| **Individual-level** |  |
| **Treatment non-adherence/discontinuation** |  |
| Treatment policy | 24/24 (100%) |
| **Treatment switching** |  |
| Treatment policy | 4/4 (4%) |
| **Mortality** |  |
| Hypothetical | 5/5 (100%) |
| **Not starting treatment** |  |
| Treatment policy | 10/10 (100%) |
| **Cluster-level** |  |
| **Not implementing treatment** |  |
| Treatment policy | 9/9 (100%) |
| **Treatment switching** |  |
| Treatment policy | 5/5 (100%) |
| **Treatment non-adherence/discontinuation** |  |
| Treatment policy | 29/29 (100%) |
| **By handling strategy (where inferable)** | **n/N<sup>b</sup> (%)** |
| **Individual-level intercurrent events** |  |
| Treatment policy for all IEs | 26/31 (84%) |
| Treatment policy for at least 1 IE | 29/31 (94%) |
| Hypothetical for at least 1 IE | 5/31 (16%) |
| Composite for at least 1 IE | 0 (0%) |
| **Cluster-level intercurrent events** |  |
| Treatment policy for all IEs | 30/31 (97%) |
| Treatment policy for at least 1 IE | 30/31 (97%) |
| Hypothetical for at least 1 IE | 0 (0%) |
| Composite for at least 1 IE | 1/31 (3%) |

<sup>a</sup>. Denominator relates to the number of trials in which the intercurrent event was inferable.
<sup>b</sup>. Denominator relates to the number of trials in which at least one type of intercurrent event's handling strategy was inferable.

Many trials excluded some clusters (11%) or individuals (30%) from the analysis population (Table 5). This was often done on the basis of clusters/individuals experiencing a specific type of intercurrent events such as not starting treatment or treatment discontinuation. This was a key driver for when the strategies to handle intercurrent events attribute was not inferable, as this way of handling can correspond to different estimands.^18, 19^ Reasons why the handling strategy for each type of intercurrent event was inferable or not can be found in Appendix 5 in the Supplemental Material.

**Table 5.** Summary of statistical methods used.

| Characteristics | Trials (N=73) |
| --- | --- |
| Statistical method used to analyse primary outcome |  |
| IEEs (independence estimating equations)<sup>a</sup> | 6 (8%) |
| GEEs(exch) | 3 (4%) |
| Mixed-effects models<sup>d</sup> | 34 (47%) |
| Cluster-level summary (marginal)<sup>b</sup> | 4 (5%) |
| Cluster-level summary (cluster-specific)<sup>c</sup> | 1 (1%) |
| Naïve individual level analyses (ignoring clustering) | 15 (21%) |
| Other<sup>e</sup> | 1 (1%) |
| Unclear<sup>f</sup> | 9 (12%) |
| Used a method that does not make the “no ICS” assumption (e.g., IEEs or cluster summary analysis) as an additional analysis when GEEs/mixed models were used as the main analysis | N = 37 |
| Yes | 1 (3%) |
| No | 36 (97%) |
| Adjusted for cluster size in main analysis |  |
| Yes | 1 (1%) |
| No | 72 (99%) |
| Reported number of individuals in each cluster |  |
| Yes | 3 (4%) |
| No | 70 (96%) |
| Excluded some clusters from analysis?<sup>g</sup> |  |
| Yes | 8 (11%) |
| No | 62 (85%) |
| Unclear | 3 (4%) |
| Excluded some participants from analysis?<sup>g</sup> |  |
| Yes | 22 (30%) |
| No | 45 (62%) |
| Unclear | 6 (8%) |

- a. Defined as any model that used an independence working correlation structure alongside cluster-robust SEs. - b. Defined as any analysis that is performed on cluster-level summaries to obtain an overall marginal (population average) treatment effect. - c. Defined as any analysis that is performed on cluster-level summaries to obtain a cluster-specific treatment effect. - d. Defined as any hierarchical model with a random intercept for cluster. - e. A multistate model. - f. Denotes any trial in which the model used for the statistical analysis was not reported in sufficient details to understand which model was used. - g. Any participant/cluster that was enrolled but not included in analysis.

### Statistical models

The most common statistical model was the mixed-effect model (n=34, 47%) (Table 5). IEEs were used in 6 trials (8%), GEE(exch) in 3 (4%), and analysis of cluster-level summaries in 5 (6%). For 9 trials (12%), it was unclear what estimation method was used as insufficient details were reported.

Of the 11 trials (14%) that used IEEs/cluster-level summaries, no trials clearly stated what the target estimand was (i.e., whether the target effect was individual-average or cluster-average), and therefore it was impossible to infer whether these methods aligned with the trial’s objective.

Over half of trials (n= 37, 51%) used GEEs(exch) or mixed-effect models, which rely on the assumption that there is no informative cluster size in order to be unbiased. However, only 1 trial (3%) performed an additional analysis using a method that does not rely on the assumption of non-informative cluster size (e.g., using IEEs/cluster-level summaries) as a sensitivity analysis. Only three trials (4%) reported the size of each cluster, making it difficult to assess whether informative cluster size was a possible concern.

## Discussion

### Principal findings

Despite publication of the ICH E9(R1) addendum in 2019, we found no evidence of uptake of estimands in our review of CRTs published between Oct 2023 and Jan 2024. Among the 73 trials we reviewed, no trial attempted to describe the estimand for their primary outcome.

This lack of uptake of estimands had major implications for our ability to decipher trial objectives. In over half of trials (51%), we could not infer which estimand was being targeted by the trial’s estimator based on the standard five attributes outlined in the ICH E9(R1) addendum; when including additional considerations specific to CRTs, we were unable to infer the full estimand for 79% of trials.

In addition to creating ambiguity around trial objectives, failure to specify the estimand also made it difficult to evaluate the appropriateness of the chosen statistical estimator, as well as the assumptions they made. For instance, 14% of trials used IEEs/cluster-level summary analyses without a clear indication of the target estimand, so it was impossible to determine whether the weighting scheme used was appropriate for trial objectives. Similarly, although 51% of trials used mixed-effects models or GEEs(exch), almost none used sensitivity analyses to evaluate robustness of these results to departure from the “no informative cluster size” assumption, nor reported the size of each cluster to allow readers to infer whether informative cluster size was a potential concern.

It was concerning to us how some trials handled intercurrent events in their analysis. For instance, 11% of trials excluded some clusters from the analysis, however, without further explanation or justification of this approach, it is difficult to understand which estimand is being targeted, or whether such an approach is justified. Furthermore, based on the statistical methods used, we identified that 16% of trials for which the strategy was inferable used a hypothetical strategy to handle individuals who died (i.e., estimated what the treatment effect would have been had no patients died). However, it is not clear whether investigators intended to implement this strategy, which has been criticised ^18, 19^, or if this was a simple happenstance based on the choice of analytical model.

### Implications of findings

A main driver in the ambiguity around target estimands was lack of clarity around some of the cluster-specific considerations which are not explicitly described in the ICH E9(R1) addendum. For instance, we could not infer whether investigators were interested in individual- or cluster-average effects for 63% of trials. This is because the dominant method to analyse CRTs is based on mixed-effects models or GEEs(exch), for which the implicit weighting mechanism corresponds to neither the individual-average (where individuals receiving equal weight) or the cluster-average (where clusters receiving equal weight) effect. Instead, these two estimators weight clusters by their inverse-variance, which is a function of the intraclass correlation coefficient and the cluster size.^9, 17^

This highlights that defining estimands according to the framework set out in the ICH E9(R1) addendum is not sufficient to clearly define the research question of interest in CRTs. This motivates the need for specific guidance for defining estimands in CRTs, which should include the considerations that are specific to CRTs, such as how individuals and clusters are weighted, and how cluster-level intercurrent events are handled.

This work also highlights the need for methods to increase uptake of estimands in CRTs. The CONSORT extension for CRTs was published in 2012, prior to publication of the ICH E9(R1) addendum. In any future updates of the CONSORT extension for CRTs, it would be useful to consider estimands as a potential reporting item, as this would help ensure that reports of CRTs clearly articulate their estimand, thereby allowing readers to better understand trial objectives, as well as to facilitate critical appraisal of statistical methods.

### Limitations and strengths

A limitation of the study is that only one database (MEDLINE) was searched while multiple databases are recommended for systematic reviews. However, MEDLINE has good coverage of medical journals that are likely to publish relevant CRTs ^20^, and our aim was to obtain a broad snapshot of current practice around the use of estimands in CRTs rather than comprehensively evaluate every single published CRT. Thus, the use of a single database was deemed sufficient for our objective.

The study had several strengths, including piloting of the data extraction form, as well as data extraction and checking by two independent statisticians to help minimise extraction errors.

## Conclusion

The uptake of estimands in published cluster randomised trial articles is low, making it difficult to ascertain which questions were being investigated or whether statistical estimators were appropriate for those questions. This highlights an urgent need to develop guidelines on defining estimands that cover unique aspects of CRTs to ensure clarity on research questions in these trials, as well as to consider the inclusion of estimands in any update to reporting guidelines for CRTs such as the CONSORT extension.

## Data Availability

All data produced in the present study are available upon reasonable request to the authors

## Author Contributions

DB wrote the first draft of the manuscript. AC and BCK revised the manuscript. All authors read and approved the final manuscript. The corresponding author attests that all listed authors meet authorship criteria and that no others meeting the criteria have been omitted.

## Declaration of Conflicting Interests

The Authors declare that there is no conflict of interest.

## Funding

DB, AC and BCK are funded by the UK Medical Research Council (grants MC_UU_00004/07 and MC_UU_00004/09). The funders had no role in the design and conduct of the study; collection, management, analysis, and interpretation of the data; preparation, review, or approval of the manuscript; and decision to submit the manuscript for publication.

## Notes

### Competing Interest Statement

The authors have declared no competing interest.

### Funding Statement

All authors are funded by the UK Medical Research Council (grants MC_UU_00004/07 and MC_UU_00004/09). The funders had no role in the design and conduct of the study; collection, management, analysis, and interpretation of the data; preparation, review, or approval of the manuscript; and decision to submit the manuscript for publication.

## Reference

1. ICH E9 (R1) addendum on estimands and sensitivity analysis in clinical trials to the guideline on statistical principles for clinical trials https://www.ema.europa.eu/en/documents/scientific-guideline/ich-e9-r1-addendum-estimands-sensitivity-analysis-clinical-trials-guideline-statistical-principles_en.pdf.

2. Kahan BC, Hindley J, Edwards M, et al. The estimands framework: a primer on the ICH E9(R1) addendum. BMJ 2024; 384: e076316. 20240123.

3. Li F, Tian ZZ, Bobb J, et al. Clarifying selection bias in cluster randomized trials. Clin Trials 2022; 19: 33–41.

4. Li F, Tian ZZ, Tian ZB and Li F. A note on identification of causal effects in cluster randomized trials with post-randomization selection bias. COMMUNICATIONS IN STATISTICS-THEORY AND METHODS 2024; 53: 1825–1837.

5. Kahan BC, Blette BS, Harhay MO, et al. Demystifying estimands in cluster-randomised trials. Stat Methods Med Res 2024; 33: 1211–1232.

6. Schochet PZ. Estimating average treatment effects for clustered RCTs with recruitment bias. Stat Med 2024; 43: 452–474.

7. Campbell MK, Piaggio G, Elbourne DR, et al. Consort 2010 statement: extension to cluster randomised trials. Bmj-Brit Med J 2012; 345.

8. Kahan BC, Li F, Blette B, et al. Informative cluster size in cluster-randomised trials: A case study from the TRIGGER trial. Clin Trials 2023; 20: 661–669.

9. Kahan BC, Li F, Copas AJ and Harhay MO. Estimands in cluster-randomized trials: choosing analyses that answer the right question. Int J Epidemiol 2023; 52: 107-+.

10. Wang BK, Park C, Small DS and Li F. Model-Robust and Efficient Covariate Adjustment for Cluster-Randomized Experiments. JOURNAL OF THE AMERICAN STATISTICAL ASSOCIATION 2024.

11. Hemming K and Taljaard M. Commentary: Estimands in cluster trials: Thinking carefully about the target of inference and the consequences for analysis choice. Int J Epidemiol 2023; 52: 116-118. Note.

12. Bugni F, Canay I, Shaikh A and Tabord-Meehan M. Inference for cluster randomized experiments with non-ignorable cluster sizes. arXiv preprint ArXiv:220408356 2022.

13. Leyrat C, Morgan KE, Leurent B and Kahan BC. Cluster randomized trials with a small number of clusters: which analyses should be used? Int J Epidemiol 2018; 47: 321–331.

14. Ma JH, Raina P, Beyene J and Thabane L. Comparison of population-averaged and cluster-specific models for the analysis of cluster randomized trials with missing binary outcomes: a simulation study. Bmc Med Res Methodol 2013; 13.

15. Seaman S, Pavlou M and Copas A. Review of methods for handling confounding by cluster and informative cluster size in clustered data. Stat Med 2014; 33: 5371–5387.

16. Seaman SR, Pavlou M and Copas AJ. Methods for observed-cluster inference when cluster size is informative: A review and clarifications. Biometrics 2014; 70: 449–456.

17. Wang XQ, Turner EL, Li F, et al. Two weights make a wrong: Cluster randomized trials with variable cluster sizes and heterogeneous treatment effects. Contemp Clin Trials 2022; 114.

18. Kahan BC, Morris TP, White IR, et al. Estimands in published protocols of randomised trials: urgent improvement needed. Trials 2021; 22: 686. 20211009.

19. Cro S, Kahan BC, Rehal S, et al. Evaluating how clear the questions being investigated in randomised trials are: systematic review of estimands. Bmj-Brit Med J 2022; 378.

20. Bramer WM, Giustini D and Kramer BMR. Comparing the coverage, recall, and precision of searches for 120 systematic reviews in Embase, MEDLINE, and Google Scholar: a prospective study. Systematic Reviews 2016; 5: 39.

