## Supplemental Material for "Use of estimands in cluster randomised trials: a review"

### **Appendix 1****. Full search strategy**

Search date: 23 Jan 2024

|  | **Term** | **Results** |
| --- | --- | --- |
|  | Cluster design-related items^1^ | |
| 1 | (communit* adj2 random* adj2 trial*).kw,sh,tw. | 1101 |
| 2 | (group adj2 random* adj2 trial*).kw,sh,tw. | 4909 |
| 3 | (cluster adj2 random*).kw,sh,tw. | 21852 |
| 4 | "cluster-random*".kw,sh,tw. | 19434 |
| 5 | 1 or 2 or 3 or 4 | 27569 |
|  | Terms for RCTs (Cochrane Highly Sensitive Search Strategy for identifying randomized trials in MEDLINE: sensitivity- and precision-maximizing version (2008 revision)^2^ | |
| 6 | randomized controlled trial.pt. | 607255 |
| 7 | controlled clinical trial.pt. | 95538 |
| 8 | randomized.ab. | 632027 |
| 9 | placebo.ab. | 244999 |
| 10 | clinical trials as topic.sh. | 201632 |
| 11 | randomly.ab. | 425508 |
| 12 | trial.ti. | 301263 |
| 13 | 6 or 7 or 8 or 9 or 10 or 11 or 12 | 1574776 |
|  | Terms for excluding ineligible articles | |
| 14 | limit 13 to humans | 1257625 |
| 15 | pilot.ab,ti. | 203172 |
| 16 | crossover.ab,ti. | 75091 |
| 17 | stepped wedge.ab,ti. | 1791 |
| 18 | protocol.ti. | 83366 |
| 19 | review.pt,ti. | 3562235 |
| 20 | meta analysis.pt. | 193743 |
| 21 | feasibility.ab,ti. | 253950 |
| 22 | 15 or 16 or 17 or 18 or 19 or 20 or 21 | 4171078 |
| 23 | 5 and 14 | 19397 |
| 24 | 23 not 22 | 13664 |
| 25 | limit 24 to english language | 13430 |
| 26 | limit 25 to humans | 13430 |
| 27 | limit 26 to dt=20231001-20240115 | 192 |

### **Appendix 2. Data extraction form**

**Section 1: Trial characteristics**

| **Section 1: Trial characteristics** | **Data** |
| --- | --- |
| 1. Study ID (CRT1, CRT2…) |  |
| 2. First author last name |  |
| 3. Number of treatment arms (2, 3…) |  |
| 4. Number of clusters randomised |  |
| 5. Total sample size (no. analysed in the primary analysis) |  |
| 6. Type of intervention | -pharmacologic  -surgical  -psychosocial, behavioural, educational  -other  -multiple types |
| 7. If other, please specify (free text) |  |
| 8. Is it an equivalence or non-inferiority trial | -yes  -no  -unclear |
| 9. Primary outcome (free text) |  |
| 10. Criterion used to determine primary outcome | Criterion 1^1^  Criterion 2a^2^  Criterion 2b^3^ |

^1^If one outcome is listed as primary outcome, this will be used.

^2^If no outcomes or multiple outcomes are listed as the primary, and only one outcome used in sample size calculation, this will be used.

^3^ if no outcomes or multiple outcomes are listed as the primary, and a sample size calculation was performed for multiple outcomes or for a cluster level outcome, the first primary (or any) clinical outcome measured at participant level listed in the Objective/Outcomes section will be used.

**Section 2: Description and handling of patient-level intercurrent events.**

| **Section 2: Patient-level intercurrent events** | **Data** |
| --- | --- |
| 11. Description of **patient-level treatment nonadherence/discontinuation** | - yes  - no  - unclear |
| 12. Reason for patient-level treatment nonadherence/discontinuation, if mentioned (free text) |  |
| 13. If yes, where was it first mentioned | - part of estimand description  - part of analysis description  - part of results (reported as occurred)  - other |
| 14. If other, please specify (free text) |  |
| 15. If yes, the handling of treatment non-adherence/discontinuation | -stated  -not stated but inferable  -neither stated nor inferable |
| Explanation of reason for answer above, if applicable (free text) |  |
| 16. If stated/inferable, detail the strategy for handling | -treatment policy  -hypothetical  -composite  -while-on-treatment  -principal stratum  -other |
| 17. If other, please specify yes (free text) |  |
| Explanation of reason for answer above, if applicable (free text) |  |
| 18. Description of **patient-level treatment switching** | - yes  - no  - unclear |
| 19. Reason for patient-level treatment switching, if mentioned (free text) |  |
| 20. If yes, where was it first mentioned | - part of estimand description  - part of analysis description  - part of results (reported as occurred)  - other |
| 21. If other, please specify (free text) |  |
| 22. If yes, the handling of treatment switching | -stated  -not stated but inferable  -neither stated nor inferable |
| Explanation of reason for answer above, if applicable (free text) |  |
| 23. If stated/inferable, detail the strategy for handling | -treatment policy  -hypothetical  -composite  -while-on-treatment  -principal stratum  -other |
| 24. If other, please specify (free text) |  |
| Explanation of reason for answer above, if applicable (free text) |  |
| 25. Description of **mortality** | - yes  - no  - unclear |
| 26. Reason for mortality, if mentioned (free text) |  |
| 27. If yes, where was it first mentioned | - part of estimand description  - part of analysis description  - part of results (reported as occurred)  - other |
| 28. If other, please specify (free text) |  |
| 29. If yes, the handling of mortality | -stated  -not stated but inferable  -neither stated nor inferable |
| Explanation of reason for answer above, if applicable (free text) |  |
| 30. If stated/inferable, detail the strategy for handling | -treatment policy  -hypothetical  -composite  -while-on-treatment  -principal stratum  -other |
| 31. If other, please specify (free text) |  |
| Explanation of reason for answer above, if applicable (free text) |  |
| 32. **Description of not starting treatment** | - yes  - no  - unclear |
| 33. Reason for not starting treatment, if mentioned (free text) |  |
| 34. If yes, where was it first mentioned | - part of estimand description  - part of analysis description  - part of results (reported as occurred)  - other |
| 35. If other, please specify (free text) |  |
| 36. If yes, the handling of not starting treatment | -stated  -not stated but inferable  -neither stated nor inferable |
| Explanation of reason for answer above, if applicable (free text) |  |
| 37. If stated/inferable, detail the strategy for handling | -treatment policy  -hypothetical  -composite  -while-on-treatment  -principal stratum  -other |
| 38. If other, please specify (free text) |  |
| Explanation of reason for answer above, if applicable (free text) |  |
| 39. Description of **other patient-level intercurrent events** | - yes  - no |
| 40. **Other patient-level intercurrent event 1 (free text)** |  |
| 41. Where was it first mentioned | - part of estimand description  - part of analysis description  - part of results (reported as occurred)  - other |
| 42. If other, please specify (free text) |  |
| 43. The handling of patient-level intercurrent event 1 | -stated  -not stated but inferable  -neither stated nor inferable |
| Explanation of reason for answer above, if applicable (free text) |  |
| 44. If stated/inferable, detail the strategy for handling | -treatment policy  -hypothetical  -composite  -while-on-treatment  -principal stratum  -other |
| 45. If other, please specify (free text) |  |
| Explanation of reason for answer above, if applicable (free text) |  |
| 46. **Other patient-level intercurrent event 2 (free text)** |  |
| 47. Where was it first mentioned | - part of estimand description  - part of analysis description  - part of results (reported as occurred)  - other |
| 48. If other, please specify (free text) |  |
| 49. The handling of patient-level intercurrent event 2 | -stated  -not stated but inferable  -neither stated nor inferable |
| Explanation of reason for answer above, if applicable (free text) |  |
| 50. If stated/inferable, detail the strategy for handling | -treatment policy  -hypothetical  -composite  -while-on-treatment  -principal stratum  -other |
| 51. If other, please specify (free text) |  |
| Explanation of reason for answer above, if applicable (free text) |  |
| 52. **Other patient-level intercurrent event 3 (free text)** |  |
| 53. Where was it first mentioned | - part of estimand description  - part of analysis description  - part of results (reported as occurred)  - other |
| 54. If other, please specify (free text) |  |
| 55. The handling of patient-level intercurrent event 3 | -stated  -not stated but inferable  -neither stated nor inferable |
| Explanation of reason for answer above, if applicable (free text) |  |
| 56. If stated/inferable, detail the strategy for handling | -treatment policy  -hypothetical  -composite  -while-on-treatment  -principal stratum  -other |
| 57. If other, please specify (free text) |  |
| Explanation of reason for answer above, if applicable (free text) |  |

**Section 3: Description and handling of cluster-level intercurrent events**

| **Section 3: Cluster-level intercurrent events** | **Data** |
| --- | --- |
| 58. **Description of not implementing treatment** | - yes  - no  - unclear |
| 59. Reason for not implementing treatment, if mentioned (free text) |  |
| 60. If yes, where was it first mentioned | - part of estimand description  - part of analysis description  - part of results (reported as occurred)  - other |
| 61. If other, please specify (free text) |  |
| 62. If yes, the handling of not implementing treatment | -stated  -not stated but inferable  -neither stated nor inferable |
| Explanation of reason for answer above, if applicable (free text) |  |
| 63. If stated/inferable, detail the strategy for handling | -treatment policy  -hypothetical  -composite  -while-on-treatment  -principal stratum  -other |
| 64. If other, please specify (free text) |  |
| Explanation of reason for answer above, if applicable (free text) |  |
| 65. **Description of cluster-level treatment switching** | - yes  - no  - unclear |
| 66. Reason for cluster-level treatment switching, if mentioned (free text) |  |
| 67. If yes, where was it first mentioned | - part of estimand description  - part of analysis description  - part of results (reported as occurred)  - other |
| 68. If other, please specify (free text) |  |
| 69. If yes, the handling of treatment switching | -stated  -not stated but inferable  -neither stated nor inferable |
| Explanation of reason for answer above, if applicable (free text) |  |
| 70. If stated/inferable, detail the strategy for handling | -treatment policy  -hypothetical  -composite  -while-on-treatment  -principal stratum  -other |
| 71. If other, please specify |  |
| Explanation of reason for answer above, if applicable (free text) |  |
| 72. **Description of cluster-level treatment discontinuation** described | - yes  - no  - unclear |
| 73. Reason for cluster-level treatment discontinuation, if mentioned (free text) |  |
| 74. If yes, where was it first mentioned | - part of estimand description  - part of analysis description  - part of results (reported as occurred)  - other |
| 75. If other, please specify |  |
| 76. If yes, the handling of treatment discontinuation | -stated  -not stated but inferable  -neither stated nor inferable |
| Explanation of reason for answer above, if applicable (free text) |  |
| 77. If stated/inferable, detail the strategy for handling | -treatment policy  -hypothetical  -composite  -while-on-treatment  -principal stratum  -other |
| 78. If other, please specify |  |
| Explanation of reason for answer above, if applicable (free text) |  |
| 79. Description **of other cluster-level intercurrent events** | - yes  - no |
| 80. **Other cluster-level intercurrent event 1 (free text)** |  |
| 81. Where was it first mentioned | - part of estimand description  - part of analysis description  - part of results (reported as occurred)  - other |
| 82. If other, please specify |  |
| 83. The handling of cluster-level intercurrent event 1 | -stated  -not stated but inferable  -neither stated nor inferable |
| Explanation of reason for answer above, if applicable (free text) |  |
| 84. If stated/inferable, detail the strategy for handling | -treatment policy  -hypothetical  -composite  -while-on-treatment  -principal stratum  -other |
| 85. If other, please specify |  |
| Explanation of reason for answer above, if applicable (free text) |  |
| 86. **Other cluster-level intercurrent event 2 (free text)** |  |
| 87. Where was it first mentioned | - part of estimand description  - part of analysis description  - part of results (reported as occurred)  - other |
| 88. If other, please specify (free text) |  |
| 89. The handling of cluster-level intercurrent event 2 | -stated  -not stated but inferable  -neither stated nor inferable |
| Explanation of reason for answer above, if applicable (free text) |  |
| 90. If stated/inferable, detail the strategy for handling | -treatment policy  -hypothetical  -composite  -while-on-treatment  -principal stratum  -other |
| 91. If other, please specify (free text) |  |
| Explanation of reason for answer above, if applicable (free text) |  |
| 92. **Other cluster-level intercurrent event 3 (free text)** |  |
| 93. Where was it first mentioned | - part of estimand description  - part of analysis description  - part of results (reported as occurred)  - other |
| 94. If other, please specify (free text) |  |
| 95. The handling of cluster-level intercurrent event 3 | -stated  -not stated, but inferable  -neither stated nor inferable |
| Explanation of reason for answer above, if applicable (free text) |  |
| 96. If stated/inferable, detail the strategy for handling | -treatment policy  -hypothetical  -composite  -while-on-treatment  -principal stratum  -other |
| 97. If other, please specify (free text) |  |
| Explanation of reason for answer above, if applicable (free text) |  |

**Section 4: Primary estimand for primary outcome**

| **Section 4: Primary estimand** | **Data** |
| --- | --- |
| 98. Did the article try to describe the primary estimand? | -yes  -no  -unclear |
| Explanation of reason for answer above (free text) |  |
| 99. **Attribute – Patient population** | -stated  -not stated but inferable  -neither stated nor inferable |
| Explanation of reason for answer above (free text) |  |
| 100. **Attribute - Treatment conditions** | -stated  -not stated but inferable  -neither stated nor inferable |
| Explanation of reason for answer above (free text) |  |
| 101. **Attribute – Endpoint/variable** | -stated  -not stated but inferable  -neither stated nor inferable |
| Explanation of reason for answer above (free text) |  |
| 102. **Attribute – Population level summary measure** | -stated  -not stated but inferable  -neither stated nor inferable |
| Explanation of reason for answer above (free text) |  |
| 103. **Attribute - Handling of patient-level intercurrent event** | -stated  -not stated but inferable  -neither stated nor inferable  -not applicable |
| Explanation of reason for answer above (free text) |  |
| 104. **Additional consideration – Cluster population** | -stated  -not stated but inferable  -neither stated nor inferable |
| Explanation of reason for answer above (free text) |  |
| 105. **Additional consideration - Handling of cluster-level intercurrent event** | -stated  -not stated but inferable  -neither stated nor inferable  -not applicable |
| Explanation of reason for answer above (free text) |  |
| 106. **Additional consideration – How participants are weighted** | -stated  -not stated but inferable  -neither stated nor inferable |
| Explanation of reason for answer above (free text) |  |
| 107. If stated/inferable, what is it | - participant average (participants receive equal weight)  -cluster average (clusters receive equal weight)  -neither |
| 108. If neither, please specify (free text) |  |
| 109. **Additional consideration – How (potential) outcomes are summarised (marginal vs. cluster-specific)** | -stated  -not stated but inferable  -neither stated nor inferable |
| Explanation of reason for answer above (free text) |  |
| 110. If stated/inferable, what is it | -marginal  -cluster-specific  -neither |
| 111. If neither, please specify (free text) |  |
| 112. **Overall primary estimand with five standard attributes** | -stated  -not stated but inferable  -neither stated nor inferable |
| 113. **Overall primary estimand with all considerations** | -stated  -not stated but inferable  -neither stated nor inferable |

**Section 5: Estimation method**

| **Section 5 Estimation method** | **Data** |
| --- | --- |
| 114. Type of outcome | -binary  -continuous  -count/rate  -time to event  -other |
| 115. If other, please specify (free text) |  |
| 116. Statistical models used | -IEEs (independent estimating equation)  -GEEs (exchangeable correlation structure)  -cluster-level summary analyses (marginal)  -cluster-level summary analyses (cluster-specific)  -mixed-effects models  -naïve individual level analyses (not accounting for clustering)  -other  -unclear |
| 117. If other, please specify (free text) |  |
| 118. If unclear, please explain (free text) |  |
| 119. If IEEs are used, how was it implemented? | -GEEs (independence correlation structure)  -standard regression models with cluster-robust SEs  -other |
| 120. If other, please specify (free text) |  |
| 121. Is there any additional weighting?   (e.g., if cluster-level summaries were used, were they weighted by the no. of patients per cluster?) | -yes  -no  -unclear |
| 122. If unclear, please specify (free text) |  |
| 123. If yes, what was the analysis weighted by? | -number of patients per cluster  -inverse of number of patients per cluster  -combination of number of patients and ICC (intracluster correlation coefficient)  -inverse probability weighting  -other |
| 124. If other, please specify (free text) |  |
| 125. Did the analysis exclude any clusters post-randomisation? | -yes  -no  -unclear |
| 126. If unclear, please specify (free text) |  |
| 127. Did the analysis exclude any participants post-enrolment? | -yes  -no  -unclear |
| 128. If unclear, please specify (free text) |  |
| 129. If GEE (exchangeable correlation structure) or mixed-effects models are used, is a method which doesn’t assume no ICS (e.g., IEEs or cluster level summaries) reported in the manuscript as a secondary/supplemental analysis? | -yes  -no  -unclear |
| 130. If unclear, please specify (free text) |  |
| 131. Variation of cluster size – is the size of each cluster reported? | -yes  -no  -unclear |
| 132. If yes, please specify the following: | -maximum cluster size: _____  -minimum cluster size: _____  -ratio (max/min): ____ |
| 133. Did the analysis adjust for cluster size (was cluster size included as a covariate)? | -yes  -no  -unclear |
| 134. If unclear, please explain | Free text |

### **Appendix 3. Rules for determining the trial’s primary outcome and primary estimand**

An outcome was categorised as the primary outcome if it was listed as the primary outcome; if no outcomes or multiple outcomes were listed as the primary, the outcome used in the sample size calculation was denoted as the primary outcome; if no sample size calculation was performed or a sample size calculation was performed for multiple outcomes or for a cluster-level outcome, the first primary clinical outcome measured at individual-level listed in the Objective/Outcome section was chosen as the primary outcome.

An estimand was determined as the primary estimand if it was listed as the primary estimand; if no estimands or multiple estimands were described, the estimand corresponding to the main analysis of the primary outcome was chosen as the primary estimand. An analysis was determined as the main analysis if it was stated as the main analysis; if no analysis was designated as the main analysis, the first approach listed in the Statistical Methods section was used.

### **Appendix 4. Rules for determining whether attributes were stated, inferable, or not inferable**

An attribute was “stated” if it was explicitly described as part of an estimand statement; “inferable” if it is not stated but can be inferred from analytical approach (i.e., if based on the description of the planned analysis the estimand can be constructed); and “not inferable” if the estimand could not be constructed from the study description. The handling of intercurrent events attribute was “inferable” if handling strategies for all applicable intercurrent events were inferable.

The estimand was inferable if all relevant attributes were inferable to the authors.

### **Appendix 5. Additional results**

#### Table S1. Trial Characteristics

| **Characteristics** | **No. of trials (N=73)** |
| --- | --- |
| No. of treatment arms | |
| 2 | 63 (86%) |
| 3 | 9 (12%) |
| 4 | 1 (1%) |
| No. of clusters randomised ^a^ | |
| Median (IQR) | 25 (8, 203) |
| Total sample size ^b^ | |
| Median (IQR) | 754 (101,3414) |
| Type of intervention | |
| Pharmacologic | 3 (4%) |
| Surgical | 0 (0%) |
| Psychosocial/behavioural/educational | 65 (89%) |
| Other ^c^ | 5 (7%) |
| Non-inferiority/equivalence trial | N = 3 (4%) |
| Type of primary outcome | |
| Binary | 25 (34%) |
| Continuous | 36 (49%) |
| Count/rate | 9 (12%) |
| Time-to-event | 3 (4%) |

^a^ One trial did not report this information; denominator = 72.

^b^ Defined as the number of participants included in the primary analysis; 3 trials did not report this information; denominator = 70.

^c^ These interventions are: cleaning delivery kits, a loan to purchase water pumps and seeds and farming training, methods for sterilising nursing homes, nutritional interventions (school lunch), a package of 1-2 interns assigned to administrative roles.

#### Table S2. Reasons why handling of intercurrent events was inferable or non-inferable

| **Intercurrent event** | **Trials n/N (%)** |
| --- | --- |
| **Patient-level** | |
| **Treatment non-adherence/discontinuation** | |
| Applicable | 38/73 (52%) |
| Inferable | 24/38 (63%) |
| Reason inferable | |
| Participants with IE included in analysis, inferred as treatment policy | 24/24 (100%) |
| Not inferable | 14/38 (24%) |
| Reason not inferable | |
| Participants with IE excluded from analysis; unclear which strategy targeted | 11/14 (79%) |
| Unclear how IE handled in analysis | 3/14 (21%) |
| **Treatment switching** | |
| Applicable | 4/73 (5%) |
| Inferable | 4/4 (100%) |
| Reason not inferable | |
| Participants with IE included in analysis, inferred as treatment policy | 4/4 (100%) |
| Not inferable | 0/4 |
| **Mortality** | |
| Applicable | 19/73 (26%) |
| Inferable | 5/19 (26%) |
| Reason inferable | |
| Censored at time of death, inferred as hypothetical | 2/5 (40%) |
| Post-death outcomes imputed, inferred as hypothetical | 3/5 (60%) |
| Not inferable | 14/19 (74%) |
| Reason not inferable |  |
| Unclear how IE handled in analysis | 8/14 (57%) |
| Participants with IE excluded from analysis; unclear which strategy targeted | 6/14 (43%) |
| **Not staring treatment** | |
| Applicable | 19/73 (26%) |
| Inferable | 10/19 (53%) |
| Reasons inferable |  |
| Participants with IE included in analysis, inferred as treatment policy | 10/19 (53%) |
| Not inferable | 9/19 (47%) |
| Reason not inferable |  |
| Participants with IE exclude from analysis; unclear which strategy targeted | 9/9 (100%) |
| **Cluster-level** | |
| **Treatment non-adherence/discontinuation** | |
| Applicable | 30/73 (41%) |
| Inferable | 29/30 (97%) |
| Reason inferable |  |
| Participants with IE included in analysis, inferred as treatment policy | 29/29 (100%) |
| Not inferable | 1/30 (3%) |
| Reason not inferable |  |
| Clusters with IE excluded from analysis; unclear which strategy targeted | 1/1 (100%) |
| **Not implementing treatment** | |
| Applicable | 15/73 (21%) |
| Inferable | 9/15 (60%) |
| Reason inferable |  |
| Clusters with IE included in analysis, inferred as treatment policy | 9/9 (100%) |
| Not inferable | 6/15 (40%) |
| Reason not inferable |  |
| Clusters with IE excluded from analysis; unclear which strategy targeted | 6/6 (100%) |
| **Treatment switching** | |
| Applicable | 5/73 (7%) |
| Inferable | 5/5 (100%) |
| Reason inferable |  |
| Clusters with IE included in analysis, inferred as treatment policy | 5/5 (100%) |

#### Table S3. Other intercurrent events and reasons why the strategies for handling were inferable or non-inferable

| **Question** | n/N (%) |
| --- | --- |
| **Participant-level** | |
| **Number of trials with >= 1 other intercurrent events** | 6 |
| **Total number of other intercurrent events** | 8 |
| **Type of other intercurrent events** | |
| **Non-truncating** | |
| Receiving treatment before planned treatment initiation | 1 |
| Completed treatment before treatment was initiated | 1 |
| **Truncating** |  |
| Abortion | 2 |
| Early pregnancy loss | 1 |
| Still-birth | 1 |
| Blighted ovum | 1 |
| Developed a recorded contraindication to kidney transplant post-enrolment (which preludes the existence of outcome) | 1 |
| **Strategy to handle other intercurrent events** | |
| Stated | 0 |
| Inferable | 3/8 (38%) |
| Reason inferable |  |
| Analysis by ITT, inferred as treatment policy | 2/3 (67%) |
| Censored at time of occurrence, inferred as hypothetical | 1/3 (33%) |
| Not inferable | 5/8 (62%) |
| Reason not inferable |  |
| Participants with IEs excluded from analysis; unclear which strategy targeted | 5/5 (100%) |
| **Cluster-level** | |
| **Number of trials with >= 1 other intercurrent events** | 7 |
| **Total number of other intercurrent events** | 7 |
| **Type of other intercurrent events** | |
| Cluster-level delay in implementing intervention | 6 |
| No eligible participants in some clusters post randomisation | 1 |
| **Strategy to handle other intercurrent events** | |
| Stated | 0 |
| Inferable | 7/7 (100%) |
| Reason inferable |  |
| Analysis by ITT, inferred as treatment policy | 6/7 (86%) |
| Outcome for some clusters with no eligible participants set to 0, inferred as composite | 1/7 (14%) |
| Not inferable | 0 |

#### Table S4. Strategies for handling other intercurrent events

| **Question** |  |
| --- | --- |
| **By type of intercurrent events** | n/N^a^ (%) |
| **Participant-level (N=3)** | |
| **Receiving treatment before planned treatment initiation** | |
| Treatment policy | 1/1 (100%) |
| **Completed treatment before treatment was initiated**  1/1 | |
| Treatment policy | 1/1 (100%) |
| **Developed a recorded contraindication to kidney transplant post-enrolment (which preludes the existence of outcome)** | |
| Hypothetical | 1/1 (100%) |
| **Cluster-level (N=7)** | |
| **Cluster-level delay in implementing intervention** |  |
| Treatment policy | 6/6 (100%) |
| **No eligible participants in some clusters post randomisation** |  |
| Composite | 1/1 (100%) |
| **By strategies** | n/N^b^(%) |
| **Participant-level (N=3)** |  |
| Treatment policy for all IEs (where inferable) | 2/3 (67%) |
| Treatment policy for at least 1 IE | 2/3 (67%) |
| Hypothetical for at least 1 IE | 1/3 (33%) |
| Composite for at least 1 IE | 0 |
| **Cluster-level (N=7)** |  |
| Treatment policy for all IEs (where inferable) | 6/7 (86%) |
| Treatment policy for at least 1 IE | 6/7 (86%) |
| Hypothetical for at least 1 IE | 0 |
| Composite for at least 1 IE | 1/7 (14%) |

^a^ Denominator relates to the number of trials in which the intercurrent event was applicable.

^b^ Denominator relates to the number of trials for which the intercurrent event handling strategy was inferable

**Reference**

1.	Perez MC, Minoyan N, Ridde V, et al. Comparison of registered and published intervention fidelity assessment in cluster randomised trials of public health interventions in low- and middle-income countries: systematic review. *Trials* 2018; 19: 410. 20180731.

2.	*Cochrane Handbook for Systematic Reviews of Interventions*. 6.5 ed. 2025.
